# Prolonged dry seasons lengthen coccidioidomycosis transmission seasons: implications for a changing California

**DOI:** 10.1101/2024.10.22.24315941

**Authors:** Simon K. Camponuri, Jennifer R. Head, Philip A. Collender, Amanda K. Weaver, Alexandra K. Heaney, Kate A. Colvin, Abinash Bhattachan, Gail Sondermeyer-Cooksey, Duc J. Vugia, Seema Jain, Justin V. Remais

## Abstract

Coccidioidomycosis, a fungal disease caused by soil-borne *Coccidioides* spp., exhibits pronounced seasonal transmission, with incidence in California typically peaking in the fall. However, the influence of climate on the timing and duration of transmission seasons remains poorly understood. Using weekly data on reported coccidioidomycosis cases in California from 2000-2023, we developed a distributed-lag Markov state-transition model to estimate the effects of temperature and precipitation on the timing of transmission season onset and end. We found that transitions from cooler, wetter conditions to hotter, drier conditions accelerate season onset. Dry conditions (10^th^ percentile of precipitation) in the spring shifted season onset an average of 2.8 weeks (95% CI: 0.43-3.58) earlier compared to wet conditions (90^th^ percentile of precipitation). Conversely, transitions back to cooler, wetter conditions hastened season end, with dry fall conditions extending the season by an average of 0.69 weeks (95% CI: 0.37-1.41) compared to wet conditions. When dry conditions occurred in the spring and fall, the transmission season extended by 3.70 weeks (95% CI: 1.23-4.22). As California is expected to experience prolonged dry seasons with climate change, our findings suggest this shift may lengthen the time at which populations are at elevated coccidioidomycosis risk.

## Introduction

Coccidioidomycosis, also known as Valley fever, is a fungal infection caused by inhalation of soil-dwelling *Coccidioides* spp. Infection typically results in respiratory illness with symptoms including cough, fever, and fatigue, and in severe cases may result in meningitis or death.^1^ In recent years, coccidioidomycosis incidence has been rising across the southwestern U.S.,^2^ including in California, where incidence increased over 8-fold between 2000 (2.5 per 100,000 population)^3^ and 2021 (20.1 per 100,000 population).^4^ Coccidioidomycosis—like most infectious diseases^5–7^—exhibits seasonal trends in incidence, generally rising in the mid- to late summer, peaking in the fall and winter, and returning to baseline levels in the spring months.^8–10^ However, this pattern is variable across years, with some years displaying a seasonal rise and fall in incidence while others displaying stable high or low incidence throughout the year, and across geographies,^9^ with some regions displaying more consistent seasonal patterns than others, and the role of climate variability in structuring transmission seasons remains unclear.^8–13^ Determination of the precise onset and duration of seasonal transmission is important for effective risk communication and mitigating seasonal increases in disease incidence and associated healthcare utilization.^14,15^ Yet the precise onset and duration of—and heterogeneity in—coccidioidomycosis transmission seasons have not been estimated, even as substantial efforts have been devoted to the development of statistical methods for reliably determining seasonal characteristics for other diseases, such as influenza.^14–19^

Seasonal shifts in temperature and precipitation initiate *Coccidioides*’ transition from mycelia to arthroconidia—the latter of which is more readily aerosolized and inhaled—likely contributing to seasonal differences in human incidence rates.^8–10^ Arthroconidia form when alternating cells within the filamentous mycelia differentiate to form spores, partly in response to environmental stress such as increased temperatures and moisture deficits.^20^ What is more, arthroconidia can become aerosolized through wind erosion or soil disturbance and disperse via dust, and precipitation deficits are known to promote mineral dust and arthroconidia dispersal.^20–23^ Accordingly, hot, dry conditions in the summer have been linked to increased incidence in the fall in California,^9,10,12^ likely due to increased arthroconidia formation and aerosolization. Earlier onset of these conditions—as is expected under climate change^24^—may lead to an earlier coccidioidomycosis transmission season onset. In contrast, precipitation may remove airborne arthroconidia via wet deposition,^25^ and may limit the aerosolization of arthroconidia by increasing soil resistance to wind erosion due to higher interparticle capillary forces.^34^ Delayed onset of rainy seasons—such as is indicated by climate projections in California^24,26^—may therefore permit continued aerosolization of *Coccidioides* spores, extending the transmission season. California’s climate has exhibited substantial intra- and interannual variability in recent decades, including changing frequencies and intensities of phenomena such as droughts and atmospheric rivers that have important implications for coccidioidomycosis transmission.^10^ However, little is known about how interannual and spatial variation in meteorological conditions influence the timing and duration of coccidioidomycosis transmission seasons.

Here, we leverage fine-scale coccidioidomycosis incidence data to investigate how meteorological factors influence coccidioidomycosis season timing in California. We estimate the associations between temperature and precipitation and coccidioidomycosis season onsets and ends using a multi-state distributed-lag Markov modeling approach. We test the hypotheses that: 1) hotter, drier conditions earlier in the spring shift the onset of the coccidioidomycosis transmission season earlier; and 2) hotter, drier conditions in the fall, corresponding to a delayed onset of the rainy season prolongs the coccidioidomycosis transmission season. Improved understanding of meteorological influences on seasonal timing of *C. immitis* infections in California can strengthen public health campaigns by improving the accuracy and timing of risk communication and support projection of intra-annual incidence in the context of anticipated climate changes in California in the coming decades.^27^

## Methods

### Epidemiological, meteorological, and hydrological data

We obtained data on confirmed coccidioidomycosis cases reported in California between January 1, 2000, and April 1, 2023, from the California Department of Public Health’s reportable disease surveillance system.^3^ Since 1995, health care providers and laboratories have been required to report confirmed coccidioidomycosis cases—along with patient information, including residential address—to local public health authorities through this system. We calculated weekly incident cases at the census tract level based on estimated date of disease onset and offline geocoding of each case’s residence.^28^ Approximately 98% of cases could be geocoded to the tract level. To reduce exposure misclassification resulting from case geolocation to census tract of residence, we restricted our analysis to census tracts located in areas with more than 500 total cases and average annual incidence rates greater than 8 cases per 100,000 population. In most cases, these areas corresponded to counties, but some counties spanning divergent geographic and meteorological profiles were split into sub-counties along 500-meter elevational isoclines (Fig. 1A), as done previously.^10^ We obtained daily, 4 km gridded precipitation and temperature estimates between 2000 and 2023 from the Precipitation-elevation Regressions on Independent Slopes Model (PRISM),^29^ and calculated weekly total precipitation and average temperature for each census tract (Fig. 1C-D).

**Fig. 1.**
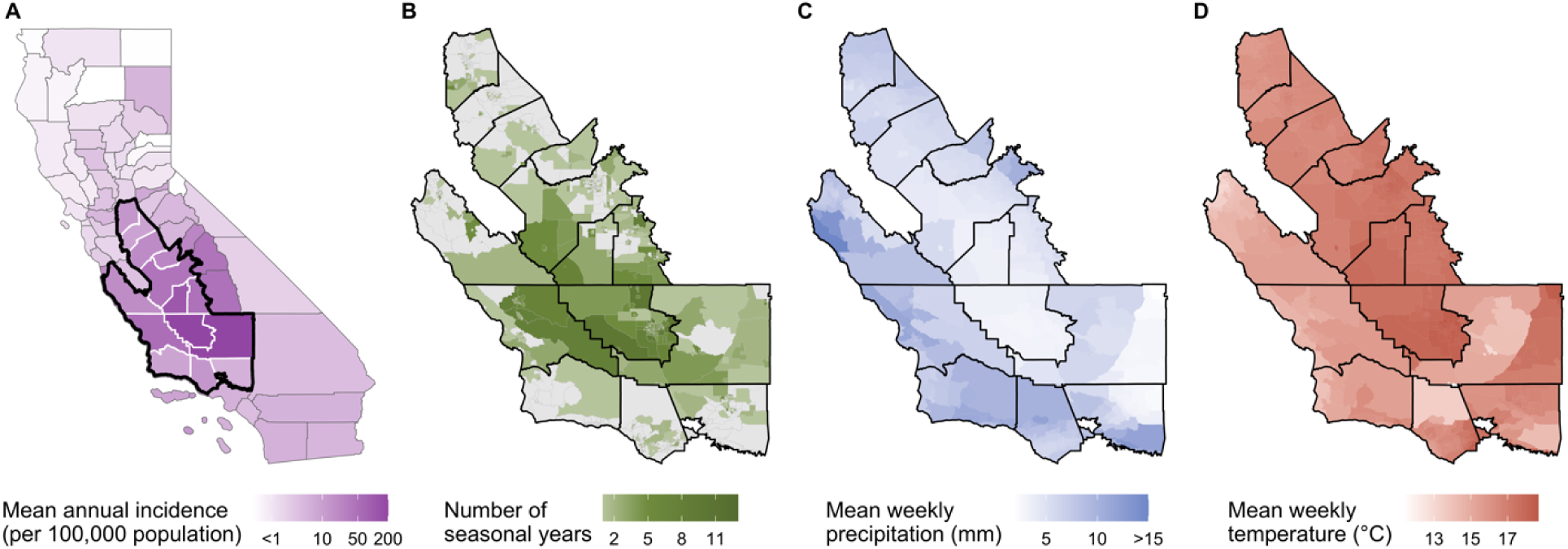
(A) County-level mean annual incidence of coccidioidomycosis between 2000-2023; counties outlined in black met the inclusion criteria (>500 cases and >8 cases per 100,000 population between 2000-2023) and sub-region designations are outlined in white. (B) Census tracts within the study area, colored by the number of transmission years exhibiting a seasonal incidence pattern over the study period; gray census tracts did not exhibit a seasonal pattern in incidence over the study period and were excluded from further analysis. (C) Average weekly precipitation (mm) over the study period among census tracts in the study area. (D) Average weekly temperature (°C) over the study period among census tracts in the study area.

### Classifying seasonal transmission years

We defined the coccidioidomycosis transmission year as spanning the average annual minima of coccidioidomycosis incidence (April 1 - March 31). To restrict our analysis to transmission years exhibiting a seasonal pattern, we first classified each census tract-transmission year as seasonal or aseasonal. To do so, we fit models of weekly incidence as a function of linear time excluding and including additional harmonic terms with 52-week periods (capturing annual seasonality between Apr. 1^st^ - Mar. 31^st^) to the incidence time-series for each census tract and transmission year. We used Akaike’s Information Criterion (AIC)^30^ to determine whether each tract-year’s data were better described by the model excluding or including harmonic terms, classifying as seasonal all tract-years where the harmonic model was preferred (Fig. 1B). We excluded from further analysis 685 census tracts (53%) where no years were classified as seasonal during the study period.

### Estimation of season onset and end

Among census tract-transmission years classified as seasonal, we estimated the week of season onset and end using segmented regression,^31,32^ which has been previously applied to influenza and other seasonal infectious diseases.^14,31,32^ The segmented regression approach involves partitioning transmission years into pre-peak and post-peak periods, then fitting piecewise linear trend lines to the incidence time-series with a single breakpoint each. The location of the breakpoint that achieves the best model fit marks the estimated onset or end of the transmission season (Fig. S1B). As the results of our analysis may be sensitive to this method of season onset and end estimation, we also estimated onset and end dates using the maximum curvature method,^14^ which estimates season onset and end as the point of maximum curvature (reciprocal of the radius of the osculating circle) in the increasing and decreasing phases of the weekly time-series, respectively (Fig. S1A), and conducted our analysis using estimates from both methods. To limit the influence of noise on season onset and end estimation, we applied a kernel smoothing function to the incidence time series with the bandwidth set to 8, as this bandwidth was the smallest value at which changes in estimates stabilized below 1 week on average compared to the next smallest bandwidth value (Fig. S2).

### Markov models of meteorological and hydrological effects on coccidioidomycosis season onset and end

To analyze the effects of temperature and precipitation on the timing of season onset and end, we modeled each census tract’s coccidioidomycosis incidence time series as a Markov process governed by two seasonal states: (1) an aseasonal state where cases are stable over time; and (2) a seasonal state where incidence increases to a seasonal peak before declining back to stable levels (Fig. 2). Using estimates of season onset and end timing (see above), we classified each week as belonging to state 1 if it occurred either before season onset or after season end and state 2 if it occurred between season onset and season end (Fig. S3). All weeks in aseasonal transmission years were classified as belonging to state 1.

**Fig. 2.**
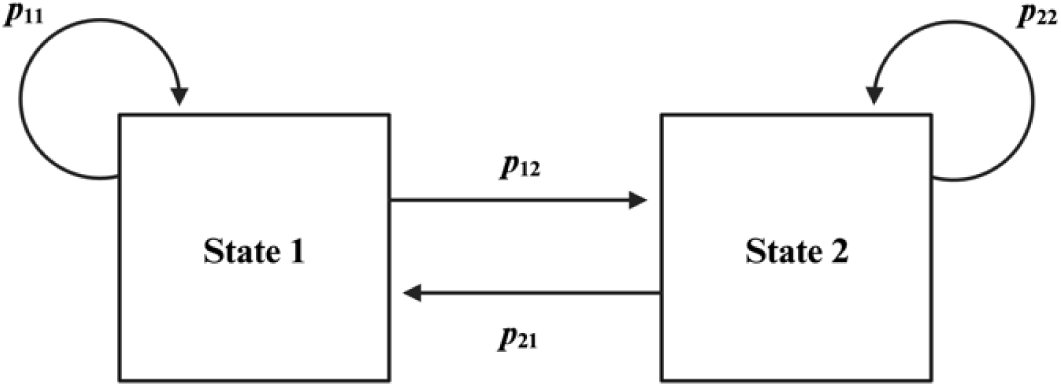
Schematic of Markov state transition process. indicates the transition from an aseasonal state to a seasonal state (i.e., season onset), and indicates the transition from a seasonal state to an aseasonal state (i.e., season end).

Per the Markov assumption, the current state of a Markov process is dependent only upon the state of the previous time step.^33^ Time-inhomogeneous Markov models relax this assumption by allowing the instantaneous probability of transition between states to vary in relation to time () and time-varying predictors ().^33^ For our analysis, transitions are governed by the transition probability matrix:

where represents the instantaneous probability of a week that was previously in state 1 staying in state 1; represents the instantaneous probability of a week that was in state 1 during the previous week transitioning to state 2; and so on. We modeled each transition probability using a proportional hazards model with the following general form:

Here, the instantaneous probability of transition from state *r* to state *S*(*P_rs_*) at time *t* is calculated as the product of the baseline transition probability (*P_Ors_*) and a log-linear function of time-varying and time-invariant covariates (***z***(***t***)) with a vector of coefficients for each pair of states (***β_rs_***). Our set of covariates included lagged temperature and precipitation as well as terms for county and year. Due to the incubation period of coccidioidomycosis (1-3 weeks)^34^ and delays in case reporting,^35,36^ we anticipated individuals to be exposed 1-12 weeks before appearing in the surveillance record. Thus, to estimate the immediate and delayed effects of temperature and precipitation on season onset and end, we used a distributed-lag non-linear (DLNM) modeling approach to estimate the effect of temperature and precipitation lagged by 2-26 weeks.^37^ Our model of season onset and end took the following form for *z*(*t*) in equation [2]:

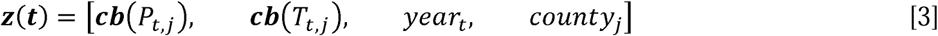

where the primary environmental exposures of interest, *T_t,j_* and *P_t,j_*, are the mean temperature and total precipitation in week *t* in census tract *j*. ***cb*** indicates a matrix of cross-basis functions, in which we modeled the exposure-response relationships between lagged environmental covariates and the outcome as linear, and constrained the lag-response relationship (2-26 weeks) to follow a natural cubic spline with 3 degrees of freedom. Models were adjusted for county and year to account for secular trends across the study area and study period, respectively.

### Estimating changes in seasonal onset and duration under counterfactual scenarios

We estimated effects of hypothetical changes in seasonal meteorological conditions on transmission season timing using G-computation substitution estimators derived from our fitted Markov models.^38,39^ To test our hypotheses, we estimated expected coccidioidomycosis season onset timing under wet vs. dry spring conditions and expected season duration under wet vs. dry fall conditions. These scenarios were selected because climate projections in California indicate a sharpening of the rainy season (i.e., reductions in both fall and spring precipitation alongside increases in winter precipitation), resulting in a prolonged dry season.^24,26^ We simulated dry and wet spring conditions by first calculating the cumulative distribution of observed spring (Mar-May) precipitation for each census tract and identifying the years in our study period where each census tract experienced the 10^th^ and 90^th^ percentile of spring precipitation. We then deterministically set the precipitation and temperature for each week to their observed values for the transmission year following a dry spring (10^th^ percentile) or a wet spring (90^th^ percentile). We simulated dry and wet fall conditions by calculating the cumulative distribution of observed fall (Sept-Nov) precipitation for each census tract and identifying the years in our study period where each census tract experienced the 10^th^ and 90^th^ percentile of fall precipitation. We then deterministically set the precipitation and temperature for each week to their observed values for the transmission year that experienced a dry fall (10^th^ percentile) or a wet fall (90^th^ percentile).

To estimate the expected time of coccidioidomycosis season onset and end 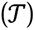 from instantaneous transition probabilities, we first calculated cumulative probabilities of transition prospectively from the start of the transmission year, for season onset, and from the estimated season onset, for season end (equation 4), with precipitation and temperature, ***A_i,t_***, and other covariates, ***w_i,t_***, deterministically set to their observed values during dry and wet conditions:

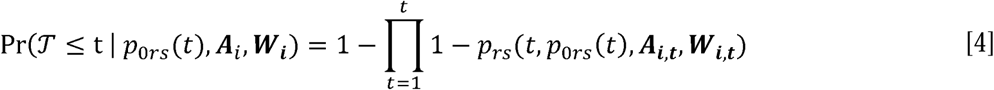

We derived distributions of census tract onset times and seasonal durations as the inverse, or quantile function, of onset and end cumulative probabilities over time. For each counterfactual contrast, we Monte Carlo simulated 10,000 onset times and durations for each condition, then empirically estimated the 2.5^th^, 50^th^, and 97.5^th^ percentiles of the distribution of the mean differences in timing between conditions of interest across census tracts.

### Sensitivity analyses

Because case data are over-dispersed such that zero cases were recorded in many weeks in many census tracts, reducing our ability to reliably detect season onset, we replicated our analysis after aggregating cases and covariates to the county (or sub-county) level and assessed the congruity of county-level and census tract-level results. To examine the sensitivity of our results to the methods of estimating season onset and end timing (i.e., SR and MC), we compared results generated using timing estimates from both methods at census tract and county (sub-county) spatial resolutions. Lastly, to assess the robustness of our results to confounding by other factors that fluctuate seasonally, we replicated our analysis after including a periodic B-spline for week in our models.

All analyses were conducted in R version 4.2.2.^40^ Piecewise linear regressions were fit using the *segmented* package,^41^ distributed-lag cross-bases were created using the *dlnm* package,^42^ and multi-state Markov models were fit using the *msm* package.^33^

## Results

### Spatial and temporal patterns of transmission season timing

Between January 1, 2000, and March 31, 2023, there were a total of 72,269 cases of coccidioidomycosis recorded in the study region. Among the counties included, western Kern County had the highest average annual incidence rate (227 per 100,000 population), while Santa Barbara County had the lowest (8.6 per 100,000 population). Only census tract-years with at least 2 cases (29.1%) exhibited a seasonal pattern, and of those, we identified 1,592 seasonal tract-years (18.5%).

On average, season onset and end occurred on weeks 17 (∼Jul 29, SD = 7.8 weeks) and 38.4 (∼Dec 26, SD = 6.9 weeks) of the transmission year (Apr. 1^st^ - Mar 31^st^), respectively, and mean season duration was 22.4 weeks (∼5.2 months, SD = 7.3). There was substantial heterogeneity in transmission season timing over the study period, with average season onsets ranging from week 10.0 (∼Jun 10) in 2002 to week 19.2 (∼Aug 14) in 2013, and season ends ranging from week 31.7 (∼Nov 9) in 2002 to week 40.8 (∼Jan 12) in 2005. Average annual season durations ranged from 17.5 weeks (∼4.0 months) long in 2000 to 24.0 weeks (∼5.5 months) long in 2005. The number of census tracts classified as seasonal varied substantially over time, with a minimum of 19 in 2000 and maximum of 206 in 2017.

Spatially, western Kern had the earliest average season onset (week 15.5 [∼Jul 19]), whereas Stanislaus had the latest (week 22.4 [∼Sep 5]; Fig. 3). Northern Los Angeles had the earliest average season end (week 35.8 [∼Dec 8]), whereas Monterey had the latest (week 42.4 [∼Jan 24]; Fig. 3).

**Fig. 3.**
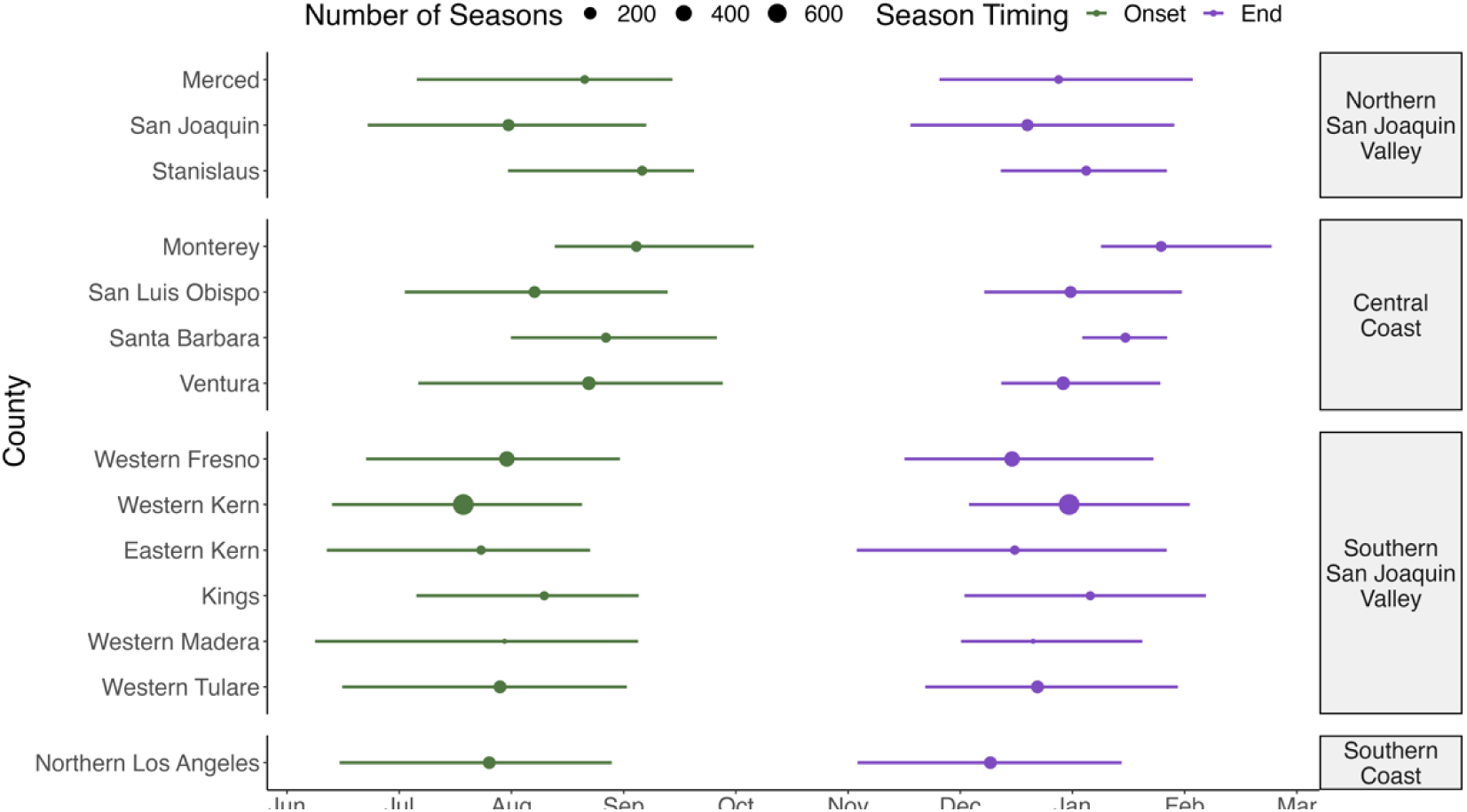
Comparison of transmission season timing across counties. Points represent average season onset (green) and end (purple) times within each county, nested within regions. Whiskers indicate the interquartile range of the estimated distributions. The size of the points corresponds to the number of seasonal tract-years within each county.

Stanislaus had the shortest average season duration (18.2 weeks [∼4.2 months]), while western Kern had the longest (24.4 weeks [5.6 months]). There was substantial spatial variation in the number of tract-years classified as seasonal across the study period, with Stanislaus having the fewest (19) and western Kern having the most (648).

### Effects of precipitation and temperature on transmission season onset

Census tract-level distributed-lag inhomogeneous Markov state models indicated that temperature and precipitation had distinct, lag-specific influences on transmission season timing (Fig. 4; Table S1). Lagged effects of increases in precipitation (adjusted for temperature) were associated with decreased onset probability at shorter lags and increased probability at longer lags. Ten mm increases in precipitation were associated with decreased onset probability at 2-14 week lags (minimum HR at 2 week lag: 0.94; 95% CI: 0.91-0.97) and increased onset probability at 19-26 week lags (maximum HR at 26 week lag: 1.06 (95% CI: 1.04-1.06). In other words, we estimated that a 10 mm increase in precipitation reduced the instantaneous probability of the coccidioidomycosis season starting two weeks later by 6%. Lagged effects of temperature (adjusted for precipitation) were roughly opposite to that of precipitation; higher temperatures were associated with increased onset probability at short lags and reduced onset probability at longer lags. A one-degree Celsius increase in temperature was associated with increased onset probability at 2-9 week lags (maximum HR at 2 week lag: 1.02; 95% CI: 1.01-1.03) and decreased probability at 12-26 week lags (minimum HR at 26 week lag: 0.98; 95% CI: 0.98-0.99; Fig. 4). In other words, we estimated that a 1 °C increase in temperature increased the instantaneous probability of the coccidioidomycosis season starting two weeks later by 2%.

**Fig. 4.**
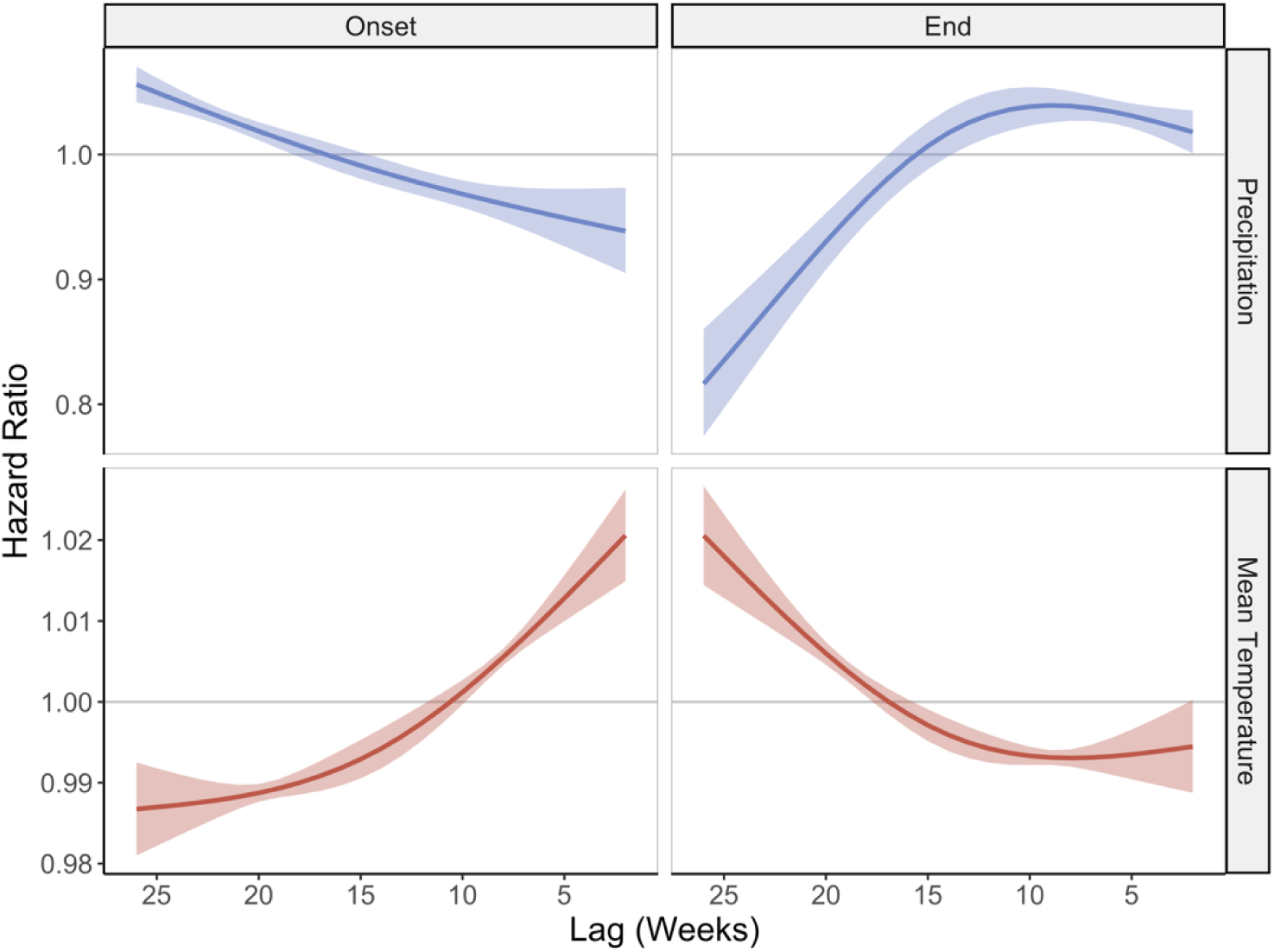
Hazard ratios associated with 10 mm increases in precipitation and 1°C increases in temperature on coccidioidomycosis season onset (left column) and end probabilities (right column) at 2-26 week lags from distributed-lag inhomogeneous Markov models. Shaded regions represent 95% confidence intervals.

Based on G-computation substitution estimators, we estimated that dry spring conditions (10^th^ percentile of precipitation) shifted the transmission season onset 2.80 weeks earlier on average (95% CI: 0.43-3.58 weeks; Fig. 5A) compared to wet spring conditions (90^th^ percentile of precipitation). Dry spring conditions reduced the probability that a given year would be seasonal; the maximum cumulative probability of season onset was lower the spring was dry (9.3%) compared to when it was wet (7.7%).

**Fig. 5.**
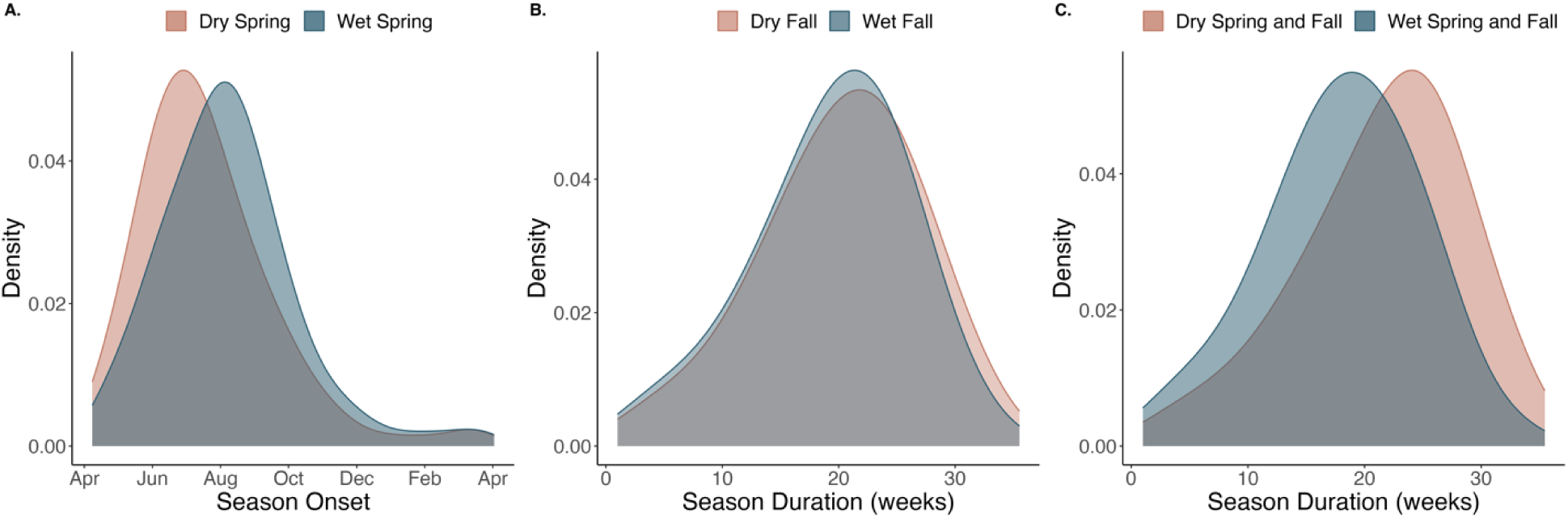
Distributions of (A) simulated season onset times given dry (brown) and wet (blue) spring conditions, (B) simulated season durations given dry (brown) and wet (blue) fall conditions, and (C) simulated season durations given both dry (brown) and wet (blue) spring and fall conditions based on G-computation substitution estimators.

### Effects of precipitation and temperature on transmission season end

For each variable, trajectories of lagged effects on transition probabilities were roughly opposite for season end compared to onset (Fig. 4). Lagged effects of increases in precipitation (adjusted for temperature) on season end were associated with increased end probability at shorter lags and decreased probability at longer lags. Ten mm increases in precipitation were associated with increased end probability at 2-13 week lags (maximum HR at 9 week lag: 1.04; 95% CI: 1.03-1.05). Conversely, at lags of 17-26 weeks, ten mm increases in precipitation were associated with reduced end probability (minimum HR at 26 week lag: 0.82, 95% CI: 0.77-0.86). Lagged effects of temperature (adjusted for precipitation) were roughly opposite; hotter temperatures were associated with reduced end probability at short lags and increased end probability at longer lags. A one-degree Celsius increase in temperature was associated with reduced end probability at 2-15 week lags (minimum HR at 8 week lag: 0.993; 95% CI: 0.992-0.994) and increased probability at 18-26 week lags (maximum HR at 26 week lag: 1.02; 95% CI: 1.01-1.03; Fig. 4).

Based on G-computation substitution estimators, we estimated that dry fall conditions (10^th^ percentile of precipitation) prolonged the coccidioidomycosis transmission season by 0.69 weeks on average (95% CI: 0.37-1.41 weeks; Fig. 5B) compared to wet fall conditions (90^th^ percentile of precipitation). Combined, dry spring and fall conditions extended the transmission season by 3.70 weeks (95% CI: 1.23-4.22 weeks) compared to wet spring and fall conditions (Fig. 5C).

### Sensitivity analyses

Seasonal transitions detected via segmented regression and maximum curvature methods were comparable, reporting concordant transition times (onset *r* = 0.70; end *r* = 0.65) (Table S3). While transmission seasons were slightly longer at the county level, with earlier onsets and later ends, estimates of mean onset and end times across counties differed by less than three weeks from census tract level estimates (Table S2). Varying the transition detection method, spatial scale, and including a periodic spline to represent seasonal effects acting through mechanisms other than temperature and precipitation resulted in similar lag-response relationships and did not qualitatively change Markov model interpretation (Figs. S4-7).

## Discussion

We found evidence that temperature and precipitation influence the timing and duration of coccidioidomycosis transmission seasons in California. Notably, shifts from cooler, wetter conditions to hotter, drier conditions—a transition typically occurring in spring—accelerated the onset of the transmission season. When the spring season was dry, the transmission season started an average of 2.8 weeks earlier compared to when the spring was wet. Conversely, shifts from hotter, drier conditions to cooler, wetter conditions—a transition typically occurring in fall—accelerated the end of the transmission season. When the fall season was dry, the transmission season was extended by an average of 0.69 weeks compared to when the fall was wet. Combined, both dry spring and fall seasons extended the length of the transmission season by a total of 3.7 weeks.

Temperature and precipitation have been previously shown to influence coccidioidomycosis incidence in California.^9–12^ Our results suggest that part of their influence on incidence may be explained by their curtailing or shifting of the coccidioidomycosis transmission season. The climate cycling associated with season onset—cool, wet conditions followed by hot, dry conditions—has been previously shown to increase coccidioidomycosis incidence,^8–10,12^ likely as a result of increased growth and subsequent dispersal of *Coccidioides* spores. Here, we build upon these studies by demonstrating that climate cycling additionally influences the timing of transmission season onset, with drier spring conditions leading to earlier increases in seasonal coccidioidomycosis transmission. Further, wet spring conditions increased the overall probability that a transmission season would occur, possibly due to increased *Coccidioides* growth in the presence of additional spring moisture, leading to a higher abundance of spores circulating during the following dry season.

The reverse climate cycle—hot, dry conditions followed by cool, wet conditions—was associated with the end of the transmission season. This result likely arises because hot, dry conditions following transmission season onset continue promoting *Coccidioides* dispersal by facilitating arthroconidia aerosolization via wind erosion and dust generation, heightening transmission. Subsequent transition to cooler, wetter conditions likely lead to a reduction in *Coccidioides* exposure, as precipitation may directly remove spores from the air^25^ or increase soil moisture and suppress dust emissions.^22^ Due to delays between pathogen exposure and disease reporting, declines in *Coccidioides* exposure may not be reflected in the coccidioidomycosis surveillance record for many weeks,^41–42^ likely why the strongest effect of precipitation occurs at a 9 week lag. Previous studies have found negative associations between concurrent precipitation and coccidioidomycosis incidence.^8–10^ Our findings concur with these and suggest that precipitation is a key factor determining when the transmission season concludes, with drier fall conditions resulting in a delayed season end and prolonged transmission of coccidioidomycosis.

Our central hypotheses were formulated based on theory for how changes in temperature and precipitation impact *Coccidioides* through their effects on the aerosol transmission efficiency of spores,^43,44^ fungal growth, and spore viability.^45,46^ However, other climate-driven processes may affect seasonal timing of *Coccidioides* transmission. For instance, *C. immitis* may be affected by soil microbial community dynamics,^47^ as fungi and bacteria in soils have been shown to have differential responses to seasonal shifts in soil moisture and temperature.^48^ Seasonality in coccidioidomycosis incidence may additionally follow from seasonal changes in human behavior or host immune defenses.^7,43^ For example, seasonal infection risk may mirror seasonal outdoor activity, including recreational (e.g., hiking^49^) and occupational (e.g., agricultural work^50^) activities that can increase exposure to ambient airborne spores as well as disturb soils (e.g., via tilling) and aerosolize the pathogen. Seasonal changes in host airway mucosal surface defenses have been associated with changes in temperature and relative humidity,^43^ which may affect the clearance of pathogens in susceptible hosts and induce seasonality in coccidioidomycosis.

Climate projections for California indicate that temperatures will continue to increase in the coming decades. These warmer temperatures are anticipated to increase moisture loss from soils, resulting in dry soil conditions that last longer into the fall and winter rainy season, even accounting for potential increases in extreme winter rain events, leading to sharper rainy seasons.^24,26,27^ According to our estimates, these changes may result in substantially longer coccidioidomycosis transmission seasons, as drier spring conditions may lead to earlier season onsets and drier fall and winter conditions may delay the end of the season. Climate change is also expected to increase the frequency of drought in the southwestern US in the 21^st^ century.^27,51^ These future droughts may disrupt the seasonal pattern of coccidioidomycosis transmission, as recent findings suggest that seasonal incidence patterns are suppressed during drought years.^13^

There are several limitations of this study. First, coccidioidomycosis cases were aggregated to census tracts based on patient home residence. This may lead to exposure misclassification and distorted seasonal timing estimates if cases were exposed to *Coccidioides* while at work or traveling in other census tracts, particularly for cases geolocated to small urban census tracts, for which movement outside of the home residence census tract is more likely. However, sensitivity analyses comparing findings at the tract level to those using a county-level spatial aggregation suggest minimal impact of small-area effects. Our seasonal transition detection methods assumed a unimodal seasonal incidence pattern. While previous work has found that coccidioidomycosis incidence in California follows a unimodal seasonal pattern on average,^7,11^ multiple seasonal peaks may be possible in some areas due to anthropogenic soil moisture alterations, such as through irrigation related to agriculture. This assumption also limits the translatability of our methods to settings where bimodal incidence patterns are observed, such as Arizona. Lastly, it is hard to disentangle seasonally varying confounders (e.g., anthropogenic activities that may impact *Coccidioides* exposure, such as agricultural or construction practices^52,53^) from seasonal variation in temperature and precipitation. Including a periodic spline to control for such seasonally varying factors attenuated the estimated effects of climate and hydrological variables in our models. However, it did not qualitatively change associations between seasonal transitions and the exposures of interest (Fig. S7).

This study also has several strengths. We investigated variation in seasonal timing of coccidioidomycosis incidence at finer spatial scales than prior work, allowing for more detailed understanding of climate factors influencing intra-annual differences in seasonality. We developed a novel Markov state transition model to estimate how spatial and temporal variation in seasonal climate patterns impact the timing and duration of coccidioidomycosis transmission seasons, leveraging a distributed-lag non-linear modeling approach to capture the effects of climate on *Coccidioides* exposure while accounting for the delays between exposure and diagnosis inherent in surveillance data.

## Conclusion

Our findings suggest that climate and hydrological factors regulate seasonal incidence patterns of coccidioidomycosis in California. By identifying how temperature and precipitation influence the onset and end of coccidioidomycosis transmission seasons, this work provides initial steps towards improved capacity to anticipate and prepare for transmission seasons based on observed or anticipated conditions. Such information could inform real-time disease surveillance efforts, public health risk assessment, and awareness campaigns targeting both the public and diagnosing physicians^54^ as well as enhance efforts to predict coccidioidomycosis incidence, particularly in the context of climate change. Future work should investigate changes to fungal growth and dust concentrations as potential mediators of the impact of climate and hydrology on coccidioidomycosis seasonal patterns to further mechanistic understanding, as well as multiannual and multi-year coccidioidomycosis incidence periodicities.

## Data Availability

The coccidioidomycosis surveillance data used here is not publicly available and is only available with explicit permission from the California Department of Public Health.

## Supplemental Tables and Figures

**Table S1.** Hazard ratios for each state transition per 1° increase in mean temperature and 10 mm increase in precipitation.

| Exposure | Lag<br>(weeks) | Hazard Ratio (95% CI) |  |
| --- | --- | --- | --- |
|  |  | Onset | End |
| Mean Temperature (°C) | 2 | 1.02 (1.01-1.03) | 0.99 (0.99-1.00) |
| Mean Temperature (°C) | 3 | 1.02 (1.01-1.02) | 0.99 (0.99-1.00) |
| Mean Temperature (°C) | 4 | 1.02 (1.01-1.02) | 0.99 (0.99-1.00) |
| Mean Temperature (°C) | 5 | 1.01 (1.01-1.02) | 0.99 (0.99-1.00) |
| Mean Temperature (°C) | 6 | 1.01 (1.01-1.01) | 0.99 (0.99-1.00) |
| Mean Temperature (°C) | 7 | 1.01 (1.01-1.01) | 0.99 (0.99-0.99) |
| Mean Temperature (°C) | 8 | 1.01 (1.00-1.01) | 0.99 (0.99-0.99) |
| Mean Temperature (°C) | 9 | 1.00 (1.00-1.00) | 0.99 (0.99-0.99) |
| Mean Temperature (°C) | 10 | 1.00 (1.00-1.00) | 0.99 (0.99-0.99) |
| Mean Temperature (°C) | 11 | 1.00 (1.00-1.00) | 0.99 (0.99-1.00) |
| Mean Temperature (°C) | 12 | 1.00 (1.00-1.00) | 0.99 (0.99-1.00) |
| Mean Temperature (°C) | 13 | 1.00 (0.99-1.00) | 0.99 (0.99-1.00) |
| Mean Temperature (°C) | 14 | 0.99 (0.99-1.00) | 1.00 (0.99-1.00) |
| Mean Temperature (°C) | 15 | 0.99 (0.99-1.00) | 1.00 (1.00-1.00) |
| Mean Temperature (°C) | 16 | 0.99 (0.99-0.99) | 1.00 (1.00-1.00) |
| Mean Temperature (°C) | 17 | 0.99 (0.99-0.99) | 1.00 (1.00-1.00) |
| Mean Temperature (°C) | 18 | 0.99 (0.99-0.99) | 1.00 (1.00-1.00) |
| Mean Temperature (°C) | 19 | 0.99 (0.99-0.99) | 1.00 (1.00-1.01) |
| Mean Temperature (°C) | 20 | 0.99 (0.99-0.99) | 1.01 (1.00-1.01) |
| Mean Temperature (°C) | 21 | 0.99 (0.99-0.99) | 1.01 (1.01-1.01) |
| Mean Temperature (°C) | 22 | 0.99 (0.99-0.99) | 1.01 (1.01-1.01) |
| Mean Temperature (°C) | 23 | 0.99 (0.98-0.99) | 1.01 (1.01-1.02) |
| Mean Temperature (°C) | 24 | 0.99 (0.98-0.99) | 1.02 (1.01-1.02) |
| Mean Temperature (°C) | 25 | 0.99 (0.98-0.99) | 1.02 (1.01-1.02) |
| Mean Temperature (°C) | 26 | 0.99 (0.98-0.99) | 1.02 (1.01-1.03) |
| Precipitation (10 mm) | 2 | 0.94 (0.91-0.97) | 1.02 (1.00-1.04) |
| Precipitation (10 mm) | 3 | 0.94 (0.91-0.97) | 1.02 (1.01-1.04) |
| Precipitation (10 mm) | 4 | 0.95 (0.92-0.97) | 1.03 (1.02-1.04) |
| Precipitation (10 mm) | 5 | 0.95 (0.93-0.97) | 1.03 (1.02-1.04) |
| Precipitation (10 mm) | 6 | 0.95 (0.93-0.97) | 1.03 (1.03-1.04) |
| Precipitation (10 mm) | 7 | 0.96 (0.94-0.97) | 1.04 (1.03-1.05) |
| Precipitation (10 mm) | 8 | 0.96 (0.95-0.97) | 1.04 (1.03-1.05) |
| Precipitation (10 mm) | 9 | 0.96 (0.95-0.98) | 1.04 (1.03-1.05) |
| Precipitation (10 mm) | 10 | 0.97 (0.96-0.98) | 1.04 (1.02-1.05) |
| Precipitation (10 mm) | 11 | 0.97 (0.96-0.98) | 1.04 (1.02-1.05) |
| Precipitation (10 mm) | 12 | 0.98 (0.97-0.99) | 1.03 (1.01-1.05) |
| Precipitation (10 mm) | 13 | 0.98 (0.97-0.99) | 1.03 (1.01-1.04) |
| Precipitation (10 mm) | 14 | 0.99 (0.98-1.00) | 1.02 (1.00-1.04) |
| Precipitation (10 mm) | 15 | 0.99 (0.98-1.00) | 1.01 (0.99-1.03) |
| Precipitation (10 mm) | 16 | 1.00 (0.99-1.01) | 0.99 (0.98-1.01) |
| Precipitation (10 mm) | 17 | 1.00 (0.99-1.01) | 0.98 (0.96-1.00) |
| Precipitation (10 mm) | 18 | 1.01 (1.00-1.02) | 0.96 (0.95-0.98) |
| Precipitation (10 mm) | 19 | 1.01 (1.00-1.02) | 0.95 (0.93-0.97) |
| Precipitation (10 mm) | 20 | 1.02 (1.01-1.03) | 0.93 (0.91-0.95) |
| Precipitation (10 mm) | 21 | 1.02 (1.02-1.03) | 0.91 (0.89-0.94) |
| Precipitation (10 mm) | 22 | 1.03 (1.02-1.04) | 0.89 (0.86-0.92) |
| Precipitation (10 mm) | 23 | 1.04 (1.03-1.04) | 0.87 (0.84-0.91) |
| Precipitation (10 mm) | 24 | 1.04 (1.03-1.05) | 0.85 (0.82-0.89) |
| Precipitation (10 mm) | 25 | 1.05 (1.04-1.06) | 0.84 (0.80-0.88) |
| Precipitation (10 mm) | 26 | 1.06 (1.04-1.07) | 0.82 (0.77-0.86) |

**Table S2.**
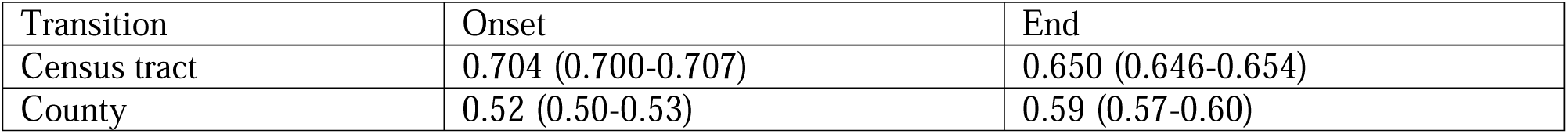
Correlation (and 95% confidence intervals) between segmented regression and maximum curvature detection method estimates at both census tract and county scales.

| Transition | Onset | End |
| --- | --- | --- |
| Census tract | 0.704 (0.700-0.707) | 0.650 (0.646-0.654) |
| County | 0.52 (0.50-0.53) | 0.59 (0.57-0.60) |

**Table S3.** Comparison of average onset, end, and duration estimates across spatial units (county vs census tract) and transition detection methods (segmented regression method and maximum curvature method).

| Scale | Method | Onset week (SD) | End week (SD) | Duration (SD) |
| --- | --- | --- | --- | --- |
| County | Segmented Regression | 15.6 (6.8) | 41.0 (6.0) | 26.4 (7.8) weeks |
| Census Tract | Segmented Regression | 17.0 (7.9) | 38.4 (7.9) | 22.4 (7.3) weeks |
| County | Maximum Curvature | 17.8 (8.7) | 40.7 (7.5) | 23.9 (9.1) weeks |
| Census Tract | Maximum Curvature | 18.1 (11.1) | 38.2 (9.4) | 21.1 (9.5) weeks |

**Fig. S1.**
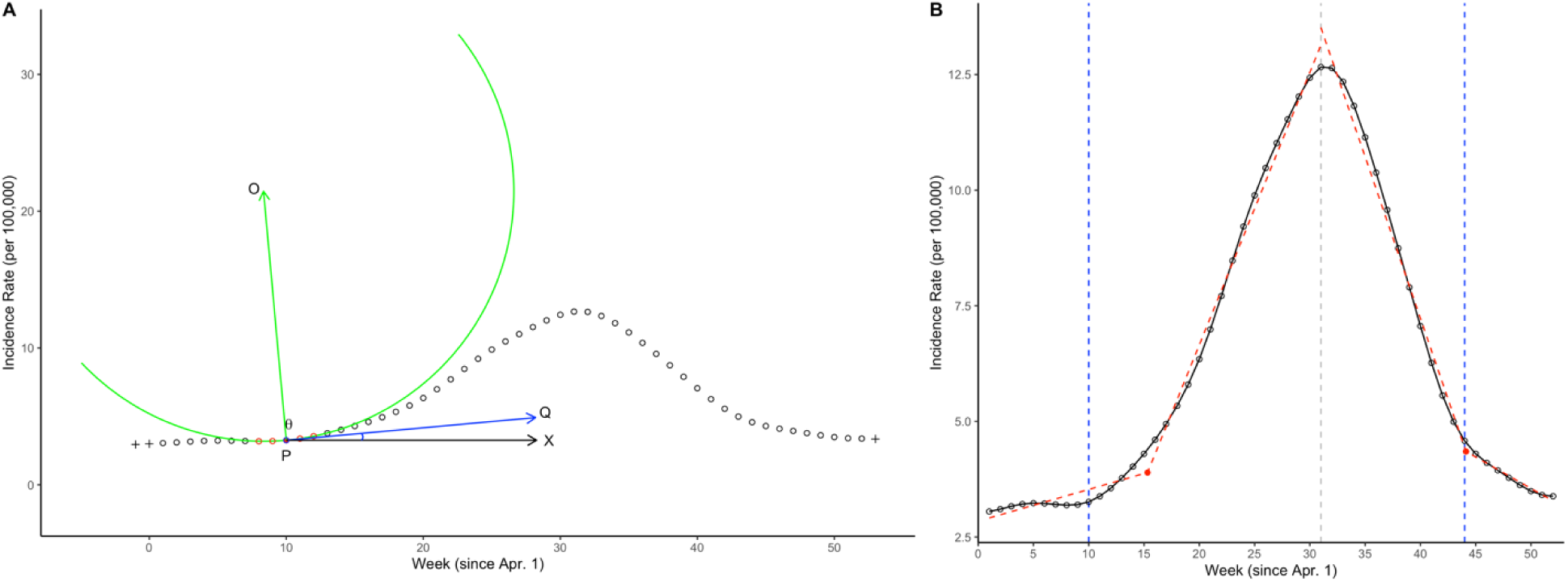
Methods used to estimate seasonal transition parameters. (A) The maximum curvature method estimates season onset and end as the point of maximum curvature (i.e., minimum angle between the tangent vector Q and the normal vector O of the osculating circle) in the increasing and decreasing phases of the weekly time-series, respectively. (B) The segmented regression method fits two piecewise linear regression lines to the incidence rate as a function of time (i.e., ordered weeks) from the beginning of the transmission year to the peak of the season (defined as the time point when the annual maximum of cases occurs), and from the peak of the season to the end of the transmission year. The breakpoint in these regression lines marks the estimated onset and end of the transmission season, respectively.

**Fig. S2.**
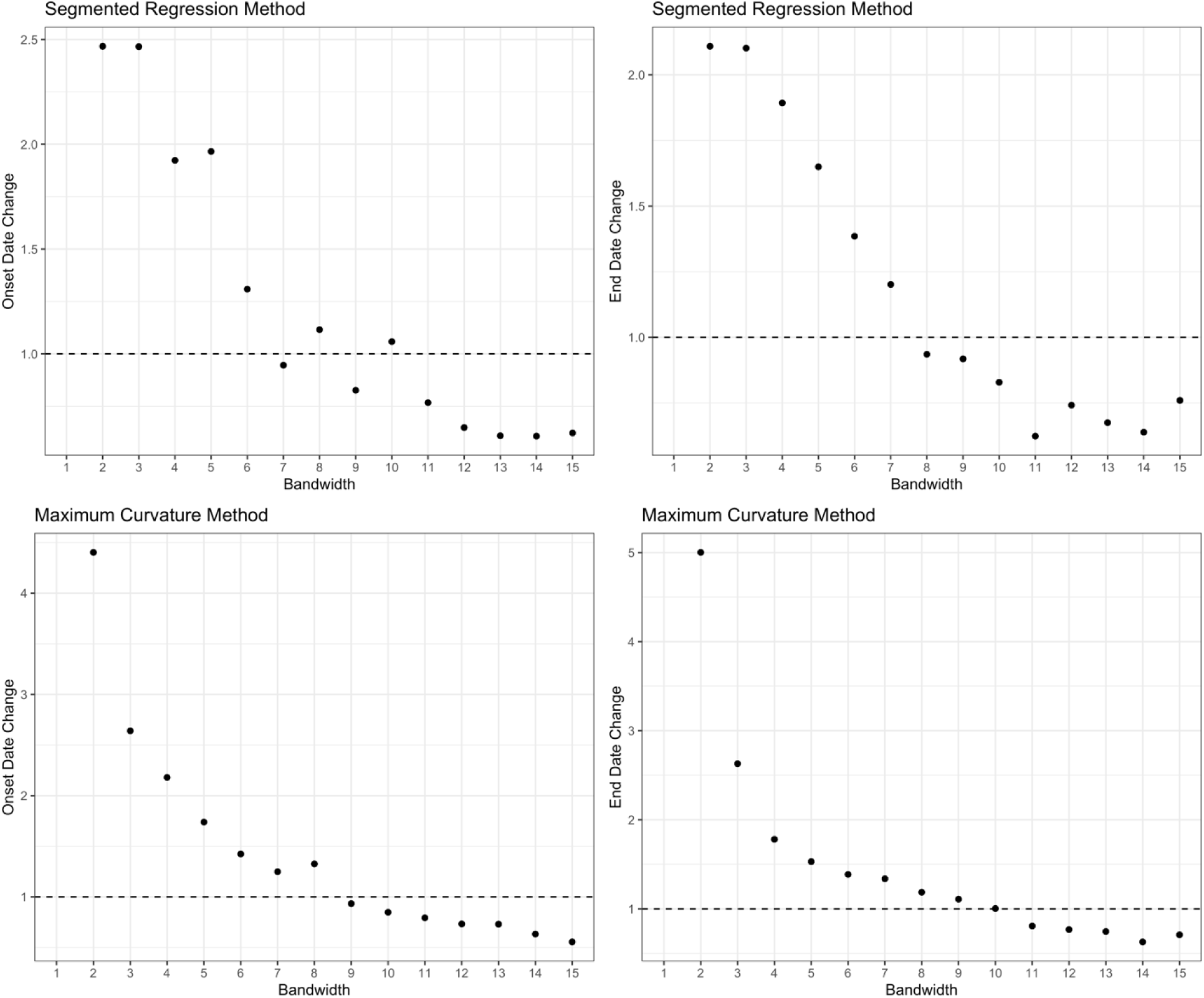
Average onset and end date estimation changes with increasing kernel smoothing bandwidth (λ). We selected a λ of 8 as it is the smallest bandwidth at which the onset and end estimates begin to stabilize below 1 week.

**Fig. S3.**
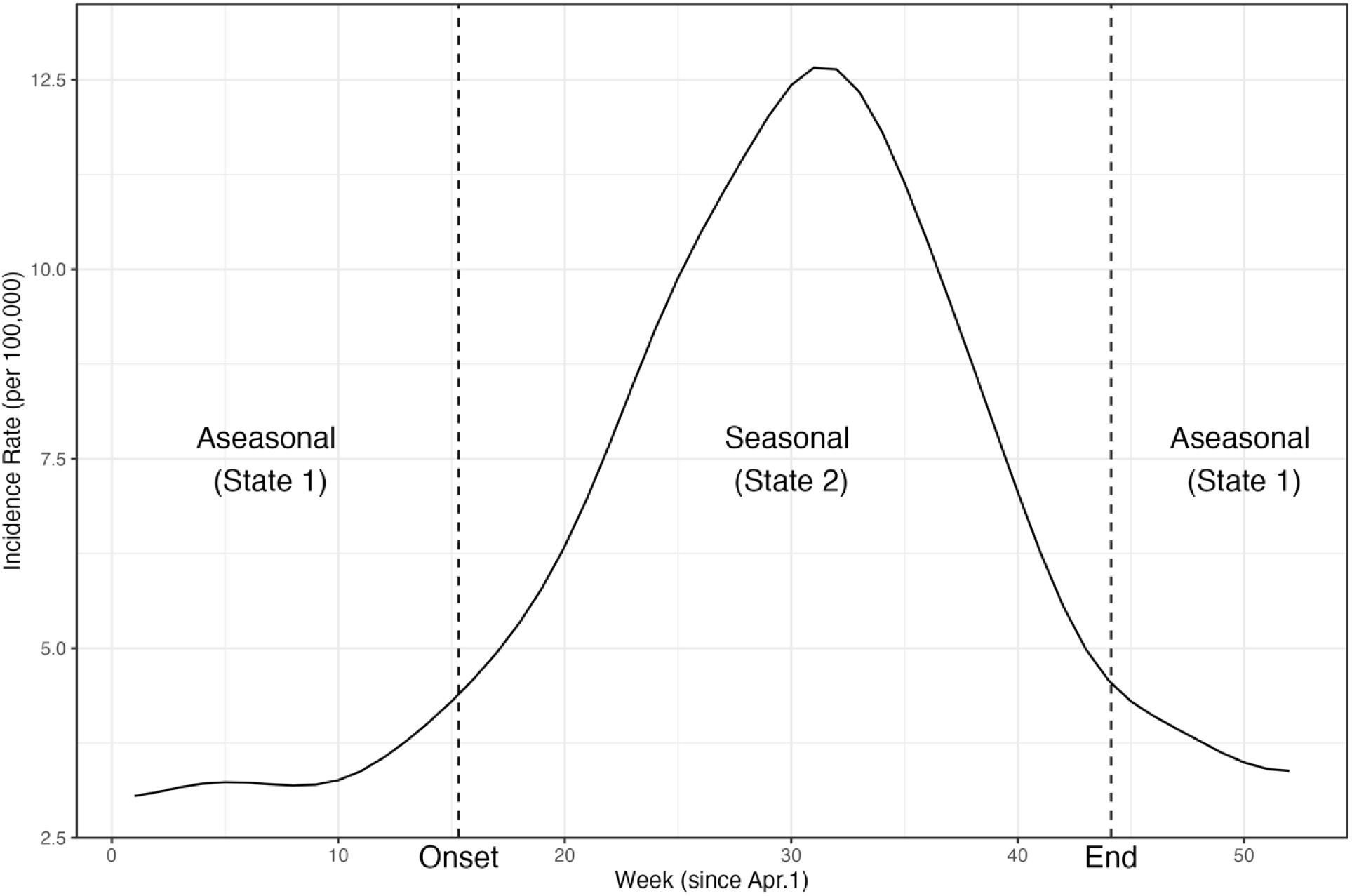
Example of the seasonal and aseasonal states (1 and 2, respectively) and transitions (onset and end). State 1 is an aseasonal state where cases are stable over time; and State 2 is a seasonal state where cases increase to a seasonal peak before declining back to a stable incidence rate. The dashed lines represent state transitions between state 1 and 2 (onset) and state 2 and 1 (end).

**Fig. S4.**
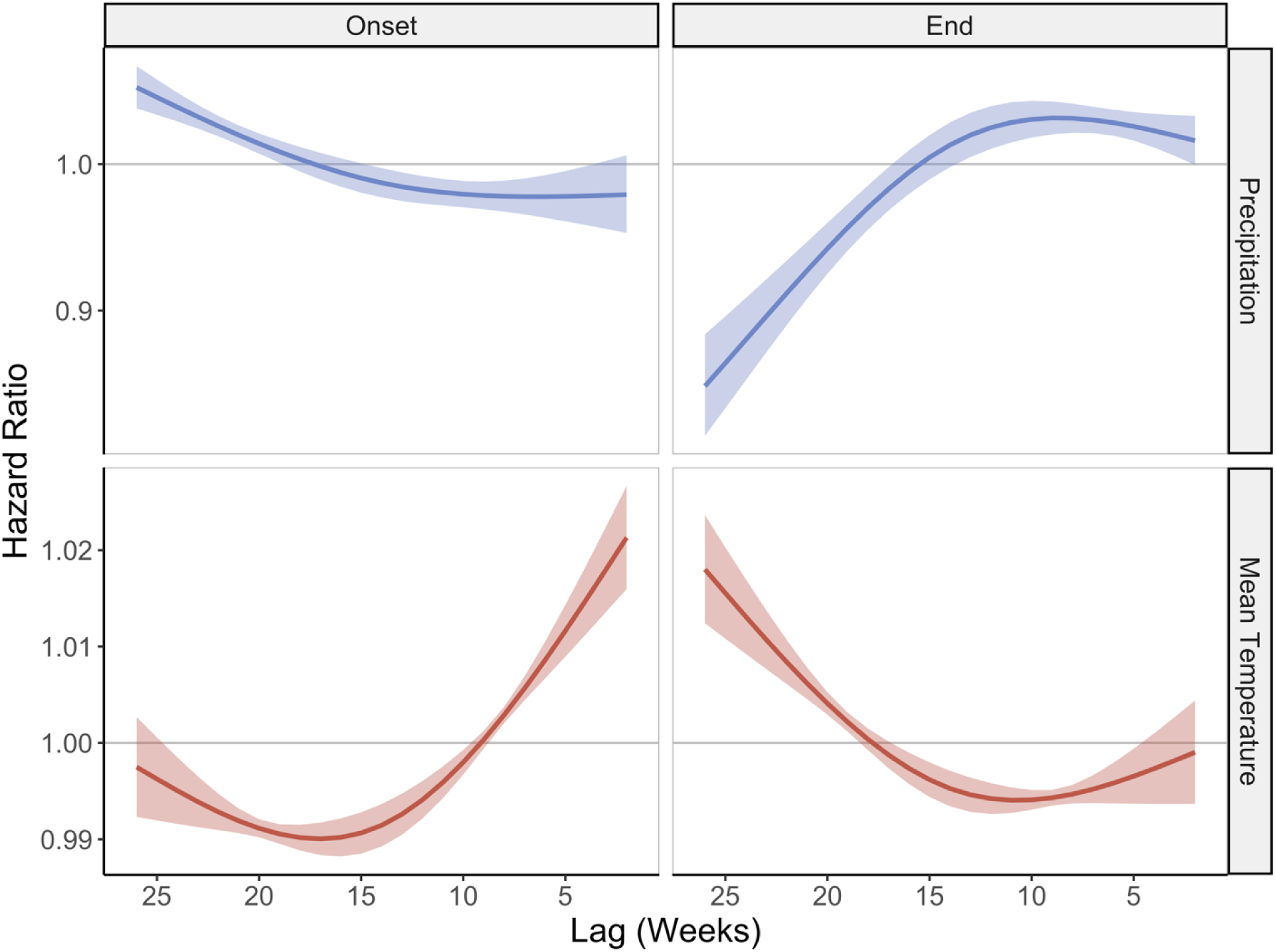
Census tract-level distributed-lag inhomogenous Markov model results using the maximum curvature detection method. Columns divide the seasonal transitions (onset and end) and rows divide the covariates (temperature and precipitation). The lines represent hazard ratios (y-axis) per 10 mm precipitation (blue) and 1 °C (red), respectively, at each week lag (x-axis) across the 2–26-week lag period associated with each week’s transition probability. The shaded regions represent the 95% confidence interval. Coefficients were constrained using natural cubic splines.

**Fig. S5.**
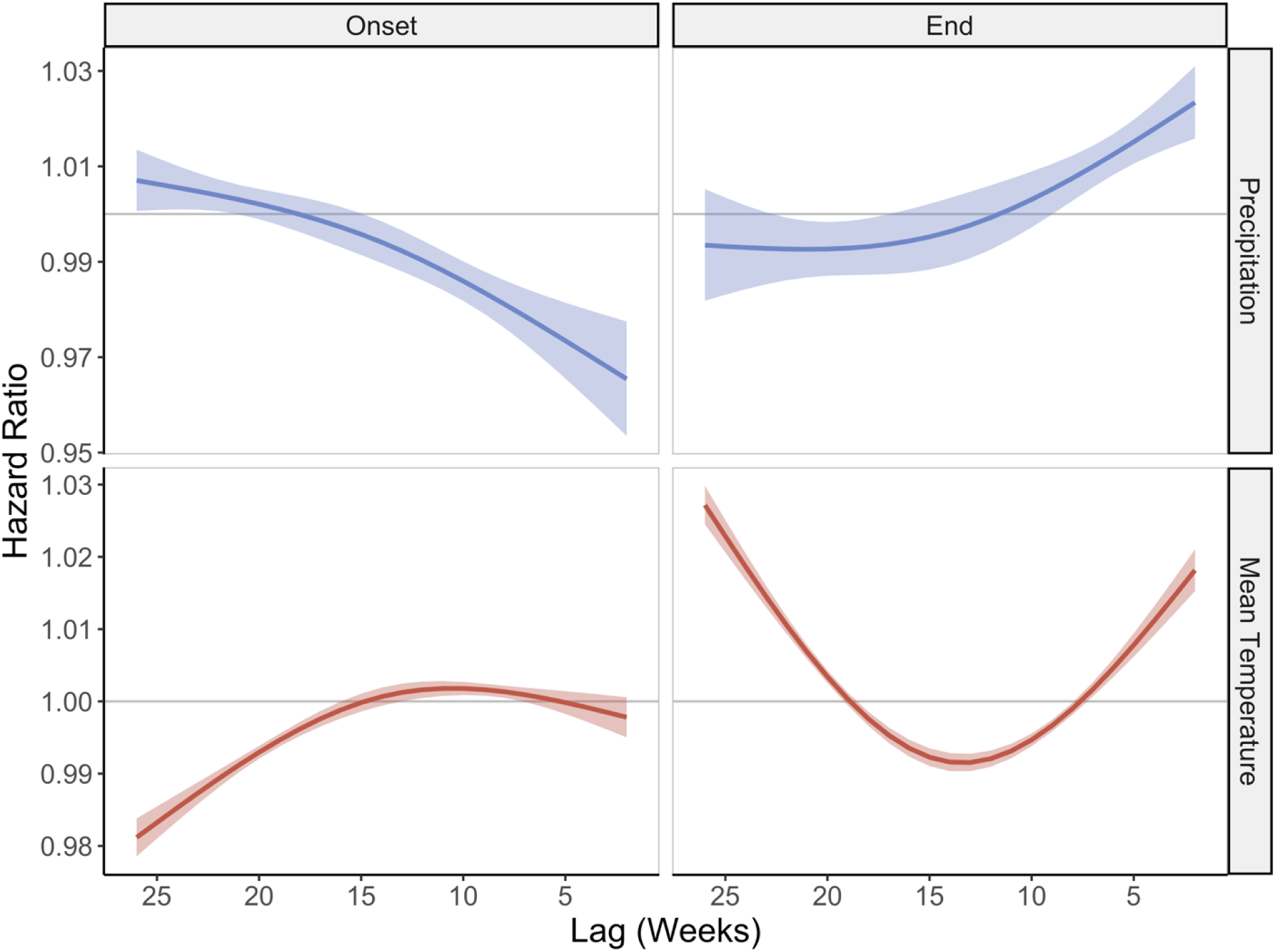
County-level distributed-lag inhomogenous Markov model results using the segmented regression detection method. Columns divide the seasonal transitions (onset and end) and rows divide the covariates (temperature and precipitation). The lines represent hazard ratios (y-axis) per 10 mm precipitation (blue) and 1 °C (red), respectively, at each week lag (x-axis) across the 2–26-week lag period associated with each week’s transition probability. The shaded regions represent the 95% confidence interval. Coefficients were constrained using natural cubic splines.

**Fig. S6.**
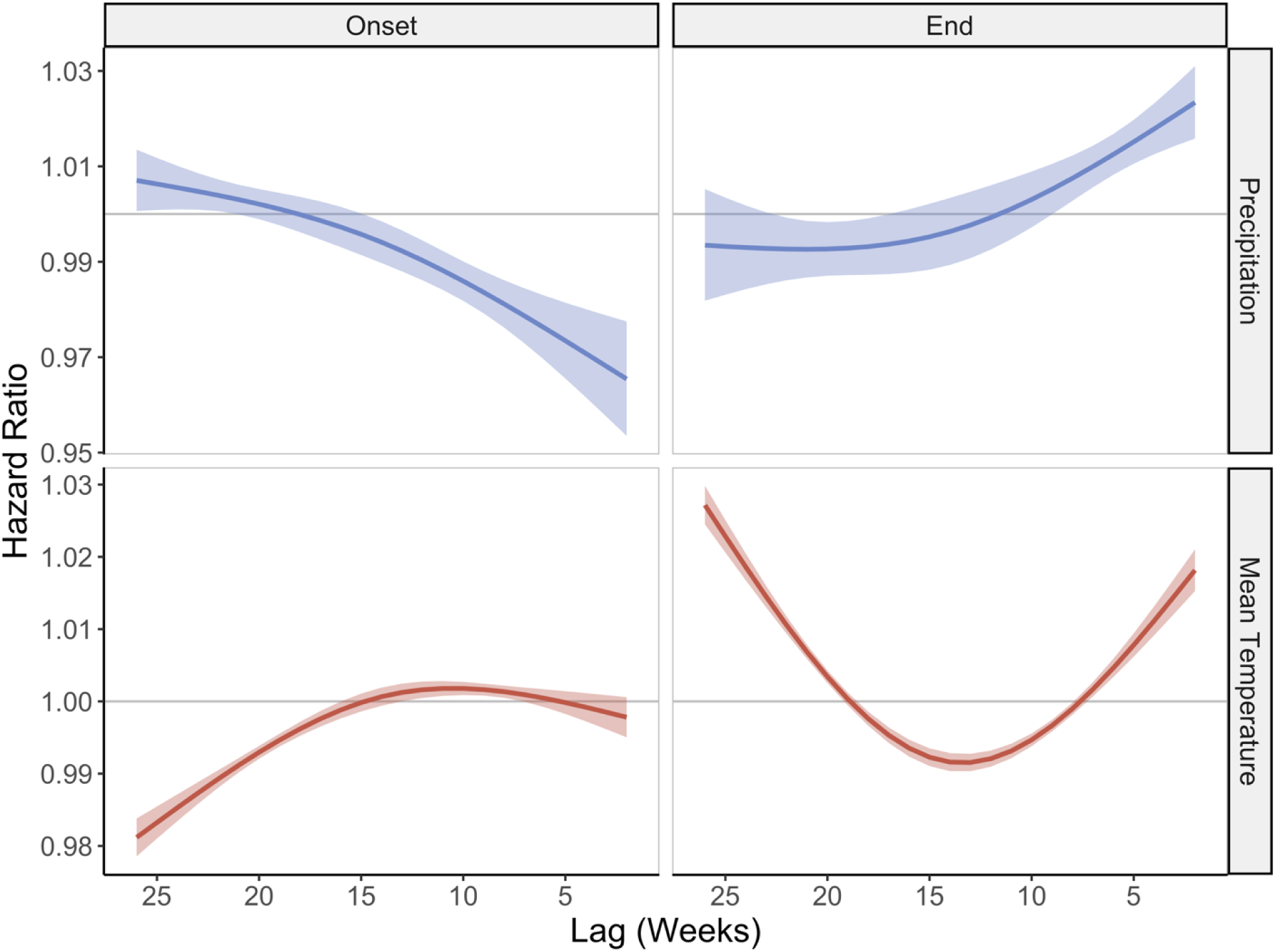
County-level distributed-lag inhomogenous Markov model results using the maximum curvature detection method. Columns divide the seasonal transitions (onset and end) and rows divide the covariates (temperature and precipitation). The lines represent hazard ratios (y-axis) per 10 mm precipitation (blue) and 1 °C (orange), respectively, at each week lag (x-axis) across the 2–26-week lag period associated with each week’s transition probability. The shaded regions represent the 95% confidence interval. Coefficients were constrained using natural cubic splines.

**Fig. S7.**
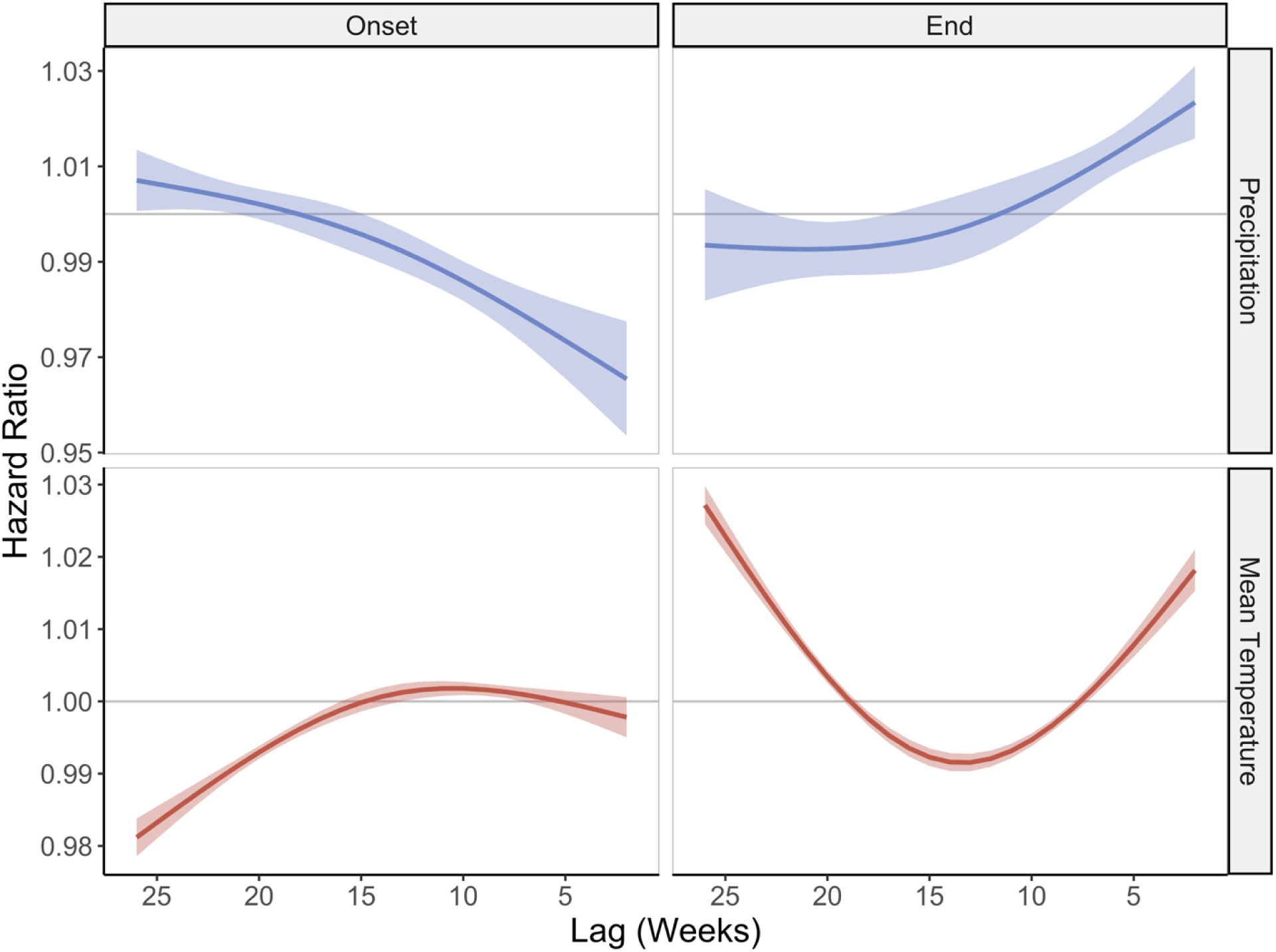
Census tract-level distributed-lag inhomogenous Markov model results using the segmented regression method, including a periodic spline on week. Columns divide the seasonal transitions (onset and end) and rows divide the covariates (temperature and precipitation). The lines represent hazard ratios (y-axis) per 10 mm precipitation (blue) and 1 °C (red), respectively, at each week lag (x-axis) across the 2–26-week lag period associated with each week’s transition probability. The shaded regions represent the 95% confidence interval. Coefficients were constrained using natural cubic splines.

